# Evaluation of Commercial Anti-SARS-CoV-2 Neutralizing Antibody Assays in seropositive subjects

**DOI:** 10.1101/2022.01.04.22268652

**Authors:** Kahina Saker, Bruno Pozzetto, Vanessa Escuret, Virginie Pitiot, Amélie Massardier-Pilonchéry, Bouchra Mokdad, Carole Langlois-Jacques, Muriel Rabilloud, Dulce Alfaiate, Nicolas Guibert, Jean-Baptiste Fassier, Antonin Bal, Sophie Trouillet-Assant, Mary-Anne Trabaud

## Abstract

The virus neutralization test (VNT) is the reference for the assessment of the functional ability of neutralizing antibodies (NAb) to block SARS-CoV-2 entry into cells. New competitive immunoassays measuring antibodies preventing interaction between the spike protein and its cellular receptor are proposed as surrogate VNT (sVNT). We tested three commercial sVNT (a qualitative immunochromatographic test and two quantitative immunoassays named YHLO and TECO) together with a conventional anti-spike IgG assay (bioMérieux) in comparison with an in-house plaque reduction neutralization test (PRNT_50_) using the original 19A strain and different variants of concern (VOC), on a panel of 306 sera from naturally-infected or vaccinated patients. The qualitative test was rapidly discarded because of poor sensitivity and specificity. Areas under the curve of YHLO and TECO assays were, respectively, 85.83 and 84.07 (p-value >0.05) using a positivity threshold of 20 for PRNT_50_, and 95.63 and 90.35 (p-value =0.02) using a threshold of 80. However, the performances of YHLO and bioMérieux were very close for both thresholds, demonstrating the absence of added value of sVNT compared to a conventional assay for the evaluation of the presence of NAb in seropositive subjects. In addition, the PRNT_50_ assay showed a reduction of NAb titers towards different VOC in comparison to the 19A strain that could not be appreciated by the commercial tests. Despite the good correlation between the anti-spike antibody titer and the titer of NAb by PRNT_50_, our results highlight the difficulty to distinguish true NAb among the anti-RBD antibodies with commercial user-friendly immunoassays.

## INTRODUCTION

Coronavirus disease 2019 (COVID-19) is an emerging disease caused by severe acute respiratory syndrome coronavirus 2 (SARS-CoV-2), and since late 2020, vaccines against SARS-CoV-2 have been available worldwide. In recent months, a large number of commercial immunoassays have been developed for the detection of specific anti-SARS-CoV-2 antibodies (1,2). However, the presence of anti-SARS-CoV-2 antibodies does not indicate whether the antibodies are able to neutralize the virus that has been reported to have a role in the protection from COVID-19 both in animals and humans (3). The gold standard for assessing the ability of antibodies to prevent the virus from entering into susceptible cells is the virus neutralization test (VNT) (4), but it requires a biosafety level 3 laboratory and takes approximately 10 days to complete. This has led to the development of SARS-CoV-2 surrogate virus neutralization tests (sVNT) that are more simple and rapid; these are based on the competition between patient antibodies and the angiotensin converting enzyme 2 (ACE2) receptor protein for binding to the spike receptor binding domain (RBD) that mediates the entry of the virus into susceptible cells (5).

These competitive immunoassays, which can be conducted using qualitative immunochromatographic cassettes or quantitative automated or manual enzyme-linked immunosorbent assay (ELISA) platforms, allow rapid and easy processing of large numbers of samples in conventional serological laboratories. However, the performance of these newly developed commercial sVNT assays by comparison to classical serological assays detecting anti-RBD IgG and/or to the reference plaque reduction neutralization test 50% (PRNT_50_) performed with live virus has been poorly evaluated up to now (6–9). Moreover, previous studies have evaluated the specificity using seronegative and/or prepandemic serum which do not inform if commercial sVNT can differentiate serum with or without neutralizing antibody in seropositive samples. The aim of the present study was to evaluate the performance of three commercial sVNT and of a classic anti-RBD IgG assay by comparison to NAb titers measured by a conventional PRNT_50_ with the original strain (clade 19A) and various clades in seropositive samples.

## MATERIALS AND METHODS

### Study

This prospective longitudinal cohort study was conducted at the laboratory associated with the national reference center for respiratory viruses (university hospital of Lyon, France). Subjects, (n=306) who were either infected with SARS-CoV-2 (n=246; 83% female; median age 41 [range: 21-66] years) or were scheduled to receive 2 doses of Pfizer BioNtech vaccine (n=31; BNT162b2/BNT162b2; 77% female; median age 41 [range: 26-69] years) or 1 dose of AstraZeneca vaccine followed by 1 dose of Pfizer BioNtech vaccine (n=29; ChAdOx1/BNT162b2; 76% female; median age 35 [range: 21-45] years) were included. For infected patients, a positive RT-PCR test was required; none of them was admitted to hospital. Blood samples were collected 6 months after infection for the convalescent cohort or 4 weeks after the two-dose vaccination for the vaccinated cohort and stored (see supplementary materials and methods).

### Serological testing

Four assays were used according to the manufacturer’s recommendations: Dynamiker Biotechnology (Tianjin, China) SARS-CoV-2 Neutralization Antibody Rapid Test, Schenzhen YHLO Biotechnologies (Schenzen, China) iFlash-2019-nCoV Nab^®^, TECO Medical Group (Sissach, Switzerland) SARS-CoV-2 Neutralization Antibody Assay, bioMérieux (Marcy l’Etoile, France) Vidas^®^ SARS-CoV-2 IgG assays. The characteristics of the assays are summarized in Table 1. For the present study suppliers kindly provided all serological kits used; there were 81 Dynamiker tests available, and a sufficient number of YHLO, TECO, and bioMérieux kits for all samples tested herein (Supplementary figure S1). YHLO and TECO are quantitative assays, bioMérieux is a semi-quantitative assay and Dynamiker is a qualitative assay. Each assay was compared to the VNT (PRNT_50_); the latter was used for the detection and titration of neutralizing antibodies, as previously described (4; see supplementary materials and methods). A threshold of 20 and of 80 was used (PRNT_50_ ≥ 20/80); the threshold of 20 of the live virus neutralization assay was considered as the detection limit of this assay. Thus, samples with PRNT_50_ titers below 20 are considered as negative for the presence of neutralizing antibodies. In contrast, the PRNT_50_ threshold of 80 was a cutoff value assumed to discriminate high from low NAb titer.

**Table 1:**
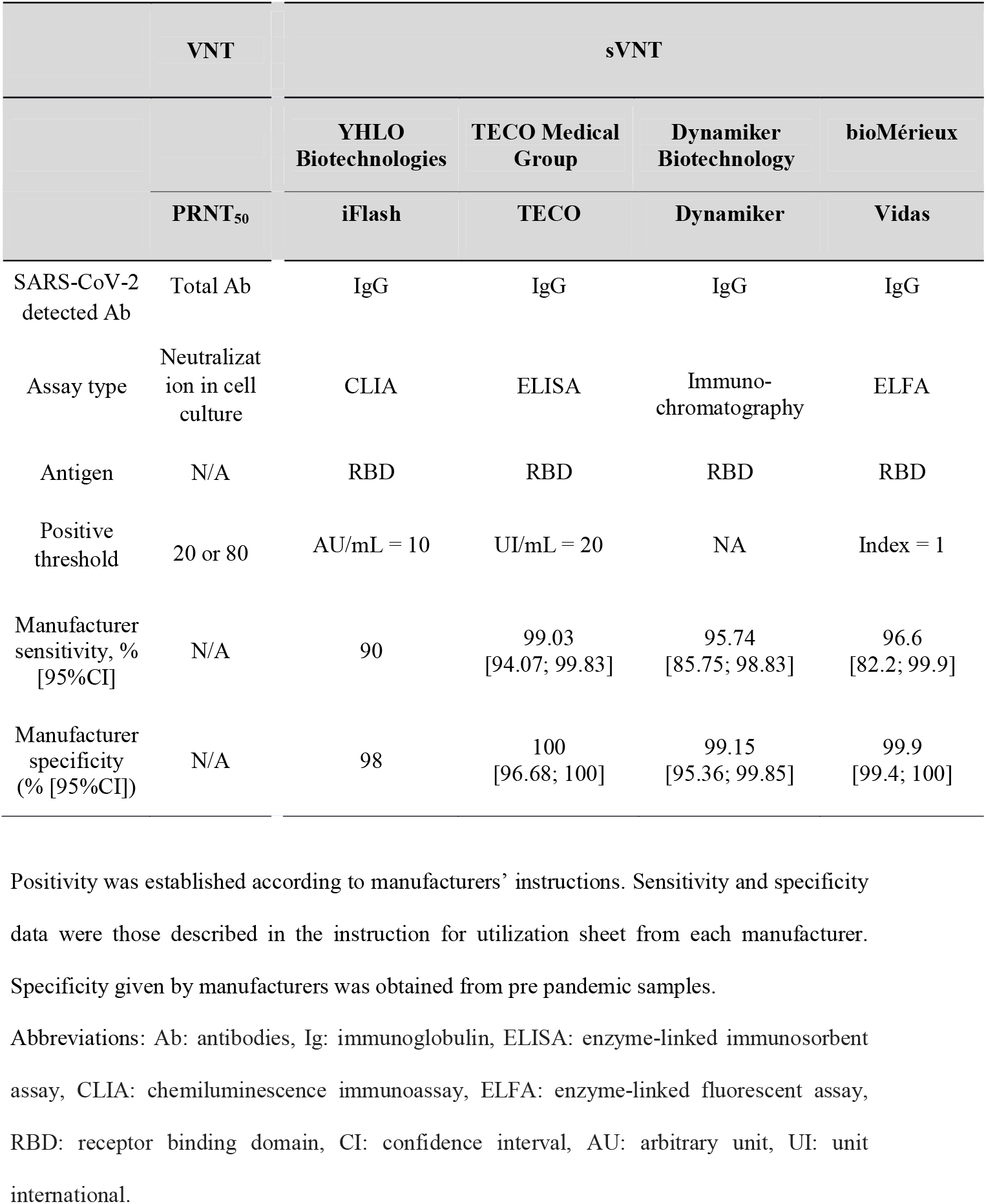
Characteristics and performance claimed by manufacturer of each assays.

First of all, we compared performance of sVNT assays with VNT (clade 19A) on 81 and 246 convalescent samples for the three sVNT and the two quantitative sVNT respectively. As the YHLO assay provided the best results among the investigated sVNT, the added-value of this test compared to a commercial serological assay detecting anti-RBD IgG (bioMérieux) was investigated considering both PRNT50 ≥ 20 and ≥80 with clade 19A, and the 246 sera from convalescent individuals and 60 sera collected 1 month post vaccination. In further experiments, we correlated the NAb titers obtained with the YHLO and bioMérieux assays to those of PRNT50 measured against various clades of SARS-CoV-2 on 60 serum specimens collected from vaccinated subjects. Each SARS-CoV-2 isolate used in this study [corresponding to 19A (B38 lineage), alpha (B.1.1.7 lineage), beta (B.1.351 lineage), gamma (P1) and delta (B.1.617.2 lineage) clades] has been sequenced to confirm the characteristic mutations of its viral clade. The sequences of the different viral strains used were deposited on Global Initiative on Sharing Avian Influenza Data (GISAID) [GISAID accession numbers: EPI_ISL_1707038 19A (B.38); EPI_ISL_1707039 Alpha (B.1.1.7); EPI_ISL_768828 Beta (B.1.351); EPI_ISL_1359892 Gamma (P.1); EPI_ISL_1904989; Delta (B.1.617.2)].

### Statistical analyses

The correlation between Ab concentrations obtained by each assay was investigated using Pearson correlation coefficients and 95% confidence interval [95%CI]. Receiver operating characteristic curves (ROC) were built to estimate the performance of YHLO, TECO, and bioMérieux assays for the detection of the presence of neutralizing antibodies considering the VNT considered as gold standard. Area under the curves (AUC) were compared using the Delong’s test. For the YHLO and TECO assays the positive threshold according to the manufacturers’ instructions were found to be too low; these were recalculated using the Youden index. Results of various clades were represented with ellipses that show the 95% CIs for different clades schedules, assuming multivariate normal distributions. Statistical analyses were conducted using GraphPad Prism^®^ software (version 8; GraphPad software, La Jolla, CA, USA) and R software, version 3.6.1 (R Foundation for Statistical Computing, Vienna, Austria). A p-value <0.05 was considered statistically significant.

### Ethics statement

Written informed consent was obtained from all participants; ethics approval was obtained from the regional review board for biomedical research in April 2020 (*Comité de Protection des Personnes Sud Méditerranée I*, Marseille, France; ID RCB 2020-A00932-37), and the study was registered on ClinicalTrials.gov (NCT04341142) (10).

## RESULTS

### Performance of sVNT

According to the VNT (PRNT_50_) with clade 19A and the positive threshold of ≥ 20, neutralizing antibodies were found in 54.5% (134/246) in the convalescent cohort and 100% (60/60) in the vaccinated cohort; using the positive threshold of ≥ 80 this was the case for 10,6% (26/246) and 95% (57/60), respectively. The performance of sVNT assays was estimated among the 81 samples for which data was available using both VNT thresholds [PRNT_50_ ≥ 20 (59/81) and PRNT_50_ ≥ 80 (24/81)] and according to the positive threshold indicated by the corresponding manufacturer. The Dynamiker qualitative test exhibited very weak performance both in terms of sensitivity and specificity, and was discarded from the following steps of the evaluation (Table 2).

**Table 2:** Performance of the three surrogate viral neutralization test compared to PRNT_50_ taken as gold standard.

|  | Dynamiker<br>Biotechnology | YHLO<br>Biotechnologies | TECO<br>Medical Group |
| --- | --- | --- | --- |
|  | Dynamiker | iFlash | TECO |
| PRNT<sub>50</sub> ≥ 20 (n=81) |  |  |  |
| Sensitivity, % [95%CI] | 49.2 [38.70; 60.10] | 100 [95.60; 100] | 98.30 [92.80; 99.60] |
| Specificity, % [95%CI] | 77.3 [60.10; 88.50] | 0 | 18.20 [8.40; 34.90] |
| PRNT<sub>50</sub> ≥ 80 (n=81) |  |  |  |
| Sensitivity, % [95%CI] | 66.7 [49.90; 80.10] | 100 [89.90; 100] | 100 [89.90; 100] |
| Specificity, % [95%CI] | 68.4 [57.70; 77.50] | 0 | 8.80 [4.30; 16.90] |
| PRNT<sub>50</sub> ≥ 20 (n=246)<br>AUC, % [IQR] | N/A | 85.83 [79.22-88.93] | 84.07 [79.22-88.93] |
| PRNT<sub>50</sub> ≥ 80 (n=246)<br>AUC, % [IQR] | N/A | 95.63 [93.11-98.15] | 90.35 [85.56-95.14] |
| Optimal threshold<br>(PRNT<sub>50</sub> ≥ 20); n=246 | N/A | 28.67 [22.58; 41.38]<br>AU/mL | 72.82 [53.85; 112]<br>UI/mL |
| Recalculated sensitivity, %<br>[95%CI] | N/A | 79.1 [62.28; 91.79] | 74.62 [59.7; 85.07] |
| Recalculated specificity, %<br>[95%CI] | N/A | 80.63 [65.18; 93.75] | 83.03 [69.64; 94.64] |
| Optimal threshold<br>(PRNT<sub>50</sub> ≥ 80; n=246) | N/A | 70.1 [53.46; 89.47]<br>AU/mL | 176.25 [124.96; 521.15]<br>UI/mL |
| Recalculated sensitivity, %<br>[95%CI] | N/A | 96.15 [92.31; 100] | 92.31 [80.76; 100] |
| Recalculated specificity, %<br>[95%CI] | N/A | 89.09 [78.63; 94.09] | 80 [70; 92.27] |

IQR: interquartile range, CI: confidence interval, AU: arbitrary unit, UI: unit international.

Despite the absence of NAbs detected using PRNT_50_ ≥ 20 in 112/246 samples and using PRNT_50_ ≥ 80 in 220/246 samples, the number of samples below the manufacturer positive threshold was 0 with the YHLO assay and 21/246 (8.5%) with the TECO assay. Among the 246 samples of convalescent individuals, the median [IQR] titers obtained using the YHLO assay this was 40.69 AU/ml [21.19-156.8] and using the TECO assay was 71.4 IU/ml [36.78-238.3]. Considering the positive threshold of 20 for PRNT_50_ as the gold standard, the AUC [95%CI] was 0.86 [0.81; 0.90] for the YHLO assay, and 0.83 [0.78; 0.88] for the TECO assay (p-value > 0.05); considering the positive threshold of 80 for PRNT_50_ as the gold standard, the AUC [95%CI] was 0.96 [0.93; 0.98] for the YHLO assay, and 0.94 [0.90; 0.97] for the TECO assay (p-value=0.02; Figure 1, Table 2). The combination of a good AUC but a very low specificity observed with the positive threshold indicated by the manufacturers led us to determine the best-fit cut-offs for the YHLO and TECO assays using the Youden index. Without weighting for the prevalence of NAb-positive samples, the median [IQR] positive threshold for the YHLO assay this was found to be 28.7 AU/ml [22.6-41.4] considering PRNT_50_ ≥ 20 as the gold standard, and 70.1 AU/ml [53.5-89.5] considering PRNT_50_ ≥ 80; for the TECO assay was found to be 72.8 IU/ml [53.9-112] considering PRNT_50_ ≥ 20 as the gold standard, and 176.3 IU/ml [125-521.2] considering PRNT_50_ ≥ 80 (Table 2). Using these optimal thresholds, considering the positive threshold of 20 for PRNT_50_ as the gold standard, the sensitivity [95%CI] and the specificity [95%CI] were 79.10 [62.28; 91.79] and 80.63 [65.18; 93.75] respectively for the YHLO assay, and 74.62 [59.7; 85.07] and 83.03 [69.64; 94.64] respectively for the TECO assay; considering the positive threshold of 80 for PRNT_50_ as the gold standard the sensitivity [95%CI] and the specificity [95%CI] were 96.15 [92.31; 100] and 89.09 [78.63; 94.09] respectively for the YHLO assay, and 92.31 [80.76; 100] and 80 [70; 92.27] respectively for the TECO assay (Table 2).

**Figure 1:**
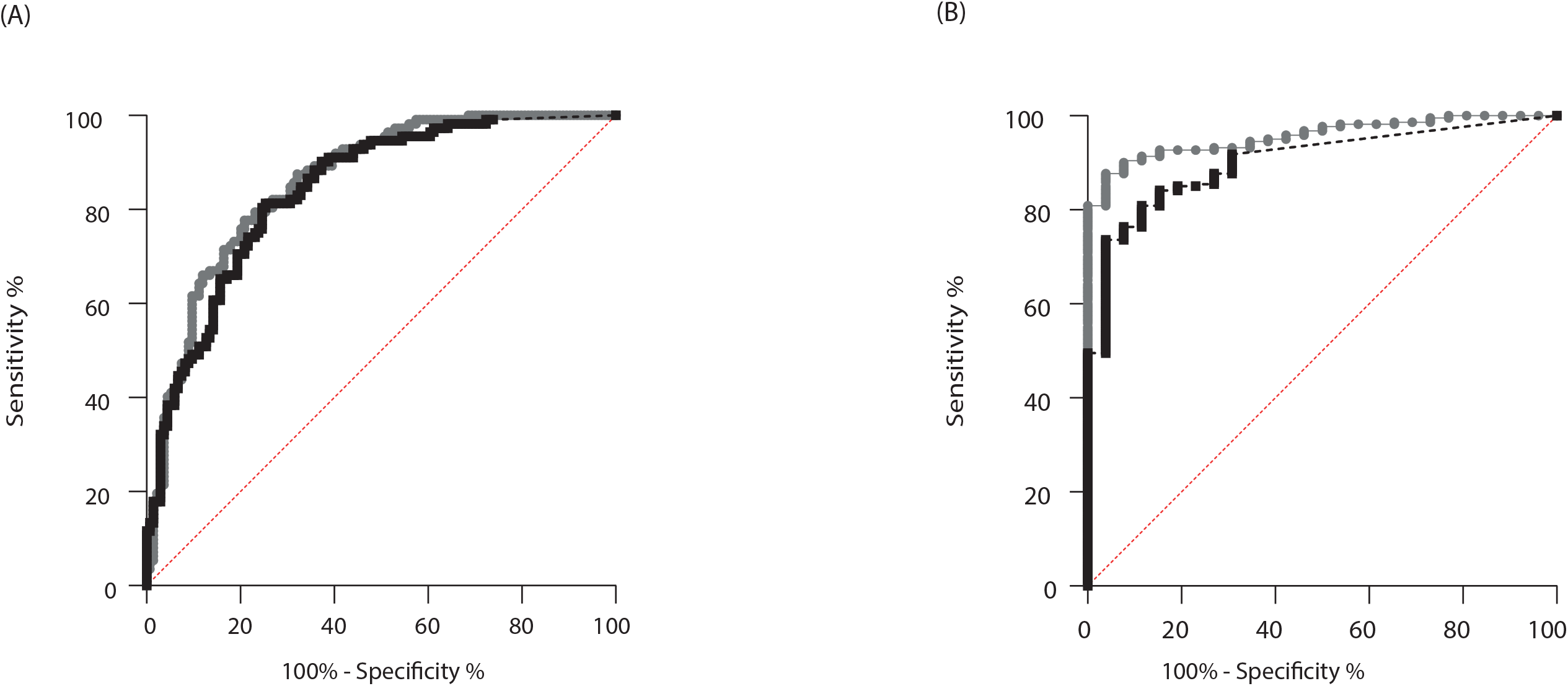
Comparison of performance of the two sVNT. ROC curves were built to estimate the performance of YHLO (in grey) and TECO (in black) assays for detecting the presence of neutralizing antibodies (PRNT_50_ ≥20 (A)) and high neutralizing antibody titre (PRNT_50_ ≥80 (B)) from samples of infected patients (n=246).

### Comparison of YHLO assay and bioMérieux anti-RBD assay with regard to PRNT_50_

The correlation coefficient (ρ [IQR]) between the YHLO assay and the VNT (PRNT_50_) was 0.85 [0.81-0.88] (Figure 2A), and between the VNT (PRNT_50_) and the bioMérieux assay it was 0.82 [0.78-0.85] (Figure 2B). Considering PRNT_50_ ≥20 as the gold standard, the AUC [IQR] for the YHLO was 0.90 [0.87-0.94] and for the bioMérieux assay it was 0.88 [0.85-0.92] (p-value > 0.05; Figure 2C); considering PRNT_50_ ≥80 as the gold standard it was 0.98 [0.96-0.99] for the YHLO assay and 0.98 [0.96-0.99] for the bioMérieux assay (p-value > 0.05; Figure 2D).

**Figure 2:**
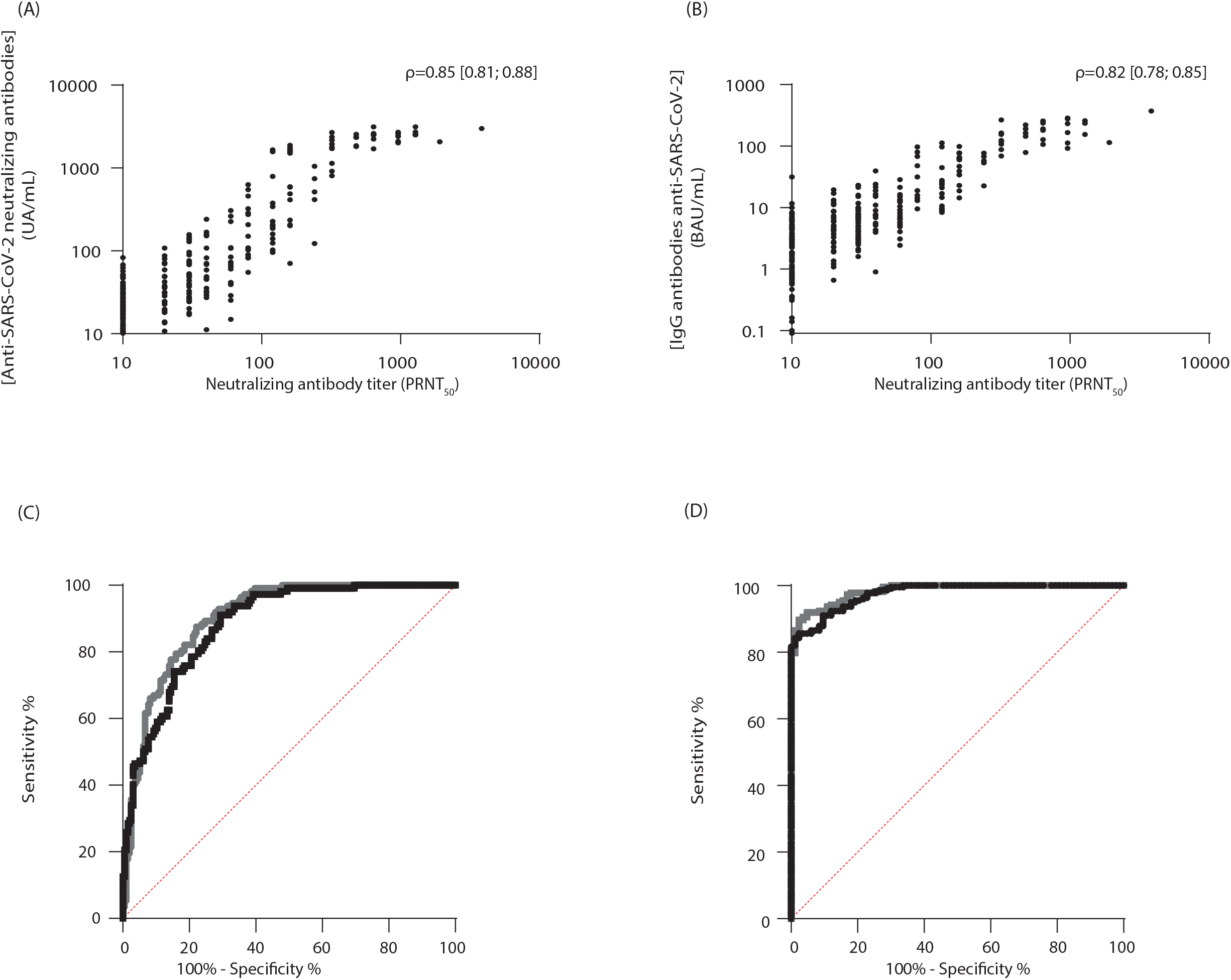
Comparison of performance of the YHLO surrogate quantitative virus neutralization test and the bioMérieux anti-RBD IgG assay with reference to the plaque reduction neutralization test 50% (PRNT_50_) from 246 serum specimens collected from convalescent patients. Panels A (YHLO assay) and B (bioMérieux assay) show the strong correlation between each test and PRNT_50_ (the value of the Spearman correlation coefficient is shown on the upper right part of the panel for each test). ROC curves were built to estimate the performance of the YHLO (in grey) and bioMérieux (in black) assays. Two different positive thresholds were used for detecting neutralizing antibodies by PRNT_50_: ≥20 (panel C) and ≥80 (panel D). The Delong test was used to compare the areas under the curve (AUC). No statistically significant difference was observed between the two tests for both thresholds.

### Impact of viral strains on Ab neutralizing capacity

Regarding the neutralizing capacity of serum against VOCs, the median fold-reduction in Nab titers varied between 1.3 against alpha strain and 2.7 against beta strain in comparison to 19A strain (Figure 3A and 3B).

**Figure 3:**
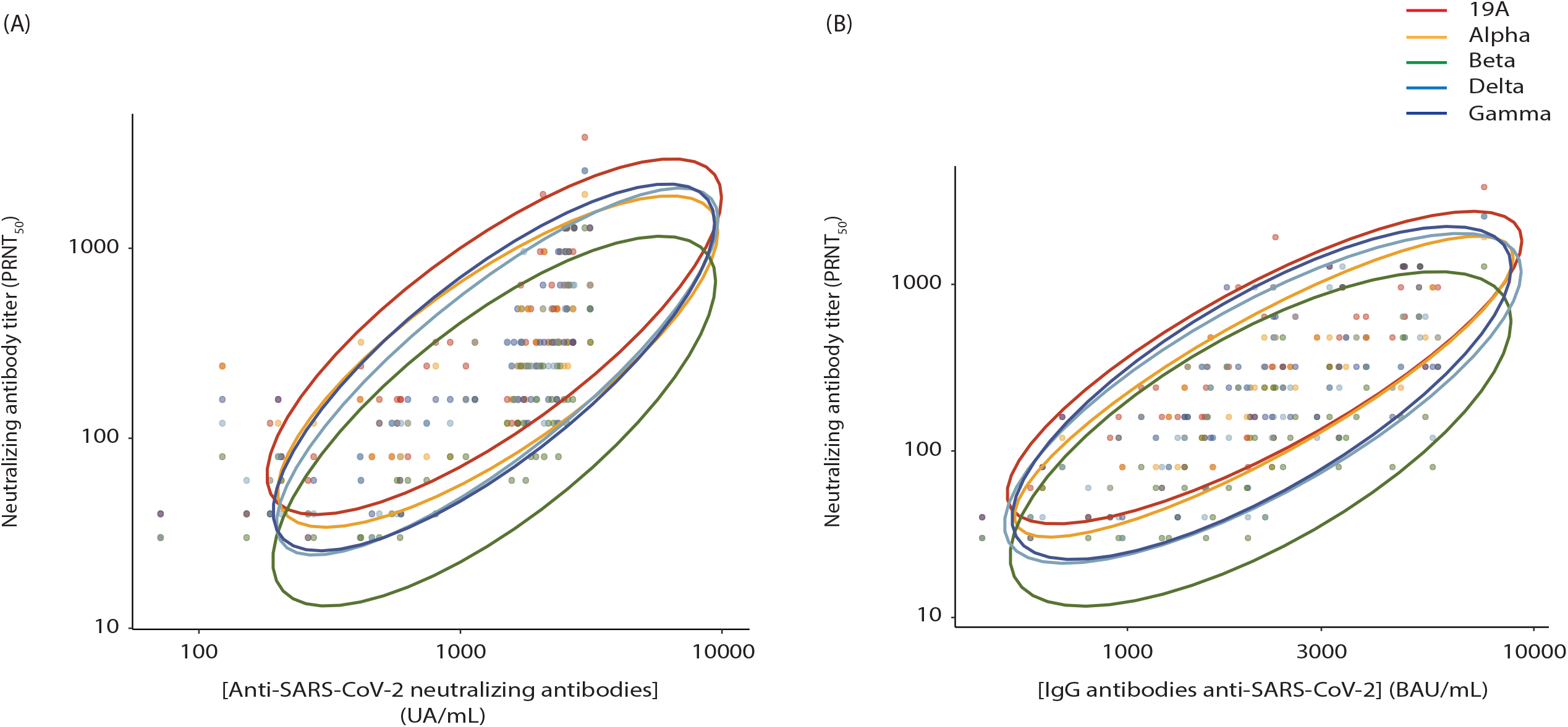
Correlation between the antibody titers obtained with the YHLO test (Panel A) or the BioMérieux assay (panel B) and the plaque reduction neutralization test 50% (PRNT50). Ellipses show the 95% CIs for different clades schedules, assuming multivariate normal distributions. Tests were performed with various clades (19A, Alpha, Beta, Gamma and Delta strains) on 60 samples taken from vaccinated individuals.

The Pearson correlation coefficient (ρ [95%CI]) for clades 19A, Alpha, Beta, Gamma and Delta: was of 0.71 [0.26; 0.91], 0.79 [0.43; 0.93], 0.71 [0.23; 0.91], 0.76 [0.36; 0.92] and 0.72 [0.29; 0.91] respectively for the YHLO assay, and of 0.86 [0.58; 0.96], 0.96 [0.86; 0.99], 0.83 [0.49; 0.95], 0.95 [0.83; 0.98] and 0.88 [0.63; 0.96] respectively for the bioMérieux assay.

Despite good correlation between concentrations of anti RBD IgG detected with YHLO or bioMérieux assays and neutralizing antibodies titers against each variant, the same titer of binding Abs overestimates titers of variant Nabs which are lower against the variants than the wild type.

## DISCUSSION

The performance of the qualitative Dynamiker assay was found to be poor, both in terms of sensitivity and in terms of specificity. The two other quantitative sVNT assays evaluated in the present study were found to be more sensitive but their specificity was extremely low since, at the manufacturers’ cutoff, most samples (TECO assay) or all of them (YHLO assay) from convalescent individuals with no detectable NAb using the live virus neutralization assay were positive for NAb with sVNT. It can be postulated that part of the antibodies detected by these ELISA are able to interfere with the interaction between ACE receptor and the viral RBD but not to prevent cell entry of the virus; this may be related to the affinity/avidity of antibodies that is reported to be low after primary infection or first vaccine dose (11,12), and is likely to be even more the case for the population included herein who were sampled 6 months after infection. Despite this low specificity, these assays correlated with the live virus neutralization assay as also found in other studies (5,6,13–15). The low specificity could also be attributed to a lack of sensitivity of live VNT but it is rather unlikely that decreasing the threshold below 20 would be clinically relevant, such low titer having little chance to be protective in vivo (16). In addition, the manufacturer positivity threshold was determined using pre-pandemic serum without any anti SARS-CoV-2 antibodies. Nevertheless, the value of sVNT use is to distinguish antibody with or without neutralizing capacity, and we designed our study to establish this performance using only seropositive samples without presence of Nab to establish specificity for sVNT assays which can explained the low specificity observed in our study.

It seems thus preferable to increase the sVNT cutoff to improve specificity, and go closer to the protective threshold. Our data from ROC curves would indicate that, for the detection of NAb, a threshold of 70 IU/ml and 30 AU/ml should be applied for the TECO an YHLO assays, respectively. However, these data have been obtained from infected subjects late after infection, at a time where antibodies are decreasing (17). This could explain the low frequency of sera with detectable NAb using the VNT, and also the discrepancy in terms of specificity between our results and previous ones using samples earlier after infection (18–21). With time, waning of antibodies could have more impact on the blocking of infection than interference with ACE binding. The study of Von Rein et al (22) suggested that the correlation between sVNT and VNT was greater at higher level of neutralization titer and they concluded that sVNT are only useful when inhibition was above 50%, which is more consistent with our data. Most of the previous studies used the cPASS assay from GeneSript, showing the correlation of competitive immunoassay to live VNT, with good sensitivity and specificity compared to VNT (5,6,9,14,15,19,20,22,23). Only a few studies reported results with the TECO (15,20) or YHLO assays (7,8,21). Of note the study of Chan et al (21) found a diagnostic cutoff, with the YHLO assay, of 27,7 AU/ml, which is closed from ours of 30 AU/ml.

Other studies have compared VNT and sVNT with assays detecting IgG binding to RBD or S proteins and observed a correlation between them (6,8,9). Fisher et al. (6) found that the correlation between sVNT and antibody binding assay is better for samples with high than low PRNT. Others studies have shown that the correlation between VNT and antibody binding assays was lower than between sVNT and antibody binding assays (9). It remains that high-throughput live virus neutralizing assays is not possible, and for this binding or competitive antibody immunoassays could be used but caution should be taken when interpreting the result, regardless of the assay used. Despite the high performance, based on the AUC of the ROC curve, of the two competitive automated immunoassays evaluated in our study, taking VNT as gold standard, we did not demonstrate any added value of sVNT compared to serological assay detecting anti-RBD IgG for evaluating the presence of Nab in seropositive subjects. These results highlight the difficulty to distinguish the Nab among anti-RBD IgG using a standard immunoassay. This difficulty could be further extrapolated considering the antibodies able to neutralize the SARS-CoV-2 variants. Using serum collected one month post full vaccination in patients with high Nab titers, we confirmed the diminution of Nab titers against different SARS-CoV-2 variant compared to initial strain. Nevertheless, the RBD coated in these competitive sVNT is not adapted to virus evolution and are not able to detect the decrease of NAb titers. To date, VNT remains the only way to detect Nabs against VOC. Taking together, from our data and those previously published, the predictive value of surrogate neutralization assays is still not obvious in all population (infected and/or vaccinated, after priming or boost immunization, early versus late after immunization). In addition, sVNT are not able to predict neutralization of variant, and thus improvements are needed before they can be considered equivalent to VNT to detect NAbs able to protect from infection.

## Data Availability

All data produced in the present study are available upon reasonable request to the authors

## ACKNOWLEDGMENTS

This study was supported by Hospices Civils de Lyon and Fondation des Hospices Civils de Lyon. The respective suppliers kindly provided all the serological kits used. We thank the staff of the occupational health and medicine department of the Hospices Civils de Lyon who contributed to the sample collection. We thank all Clinical Research Associates, for their excellent work. We thank Karima Brahima and all the members of the clinical research department for their reactivity (DRS, Hospices Civils de Lyon). Human biological samples and associated data were obtained from NeuroBioTec (CRB Hospices Civils de Lyon, Lyon France, Biobank BB-0033-00046). We thank all the technicians from the virology laboratory of the Hospices Civils de Lyon who performed the assays on the automated platforms. Lastly, we thank all the healthcare workers for their participation in this clinical study, and Philip Robinson (DRS, Hospices Civils de Lyon) for help in manuscript preparation.

---

**Supplementary figure S1: Study flow diagram.**

