## Supplementary figures and images for "Evaluation of Commercial Anti-SARS-CoV-2 Neutralizing Antibody Assays in seropositive subjects"

### Supplementary Figure S1

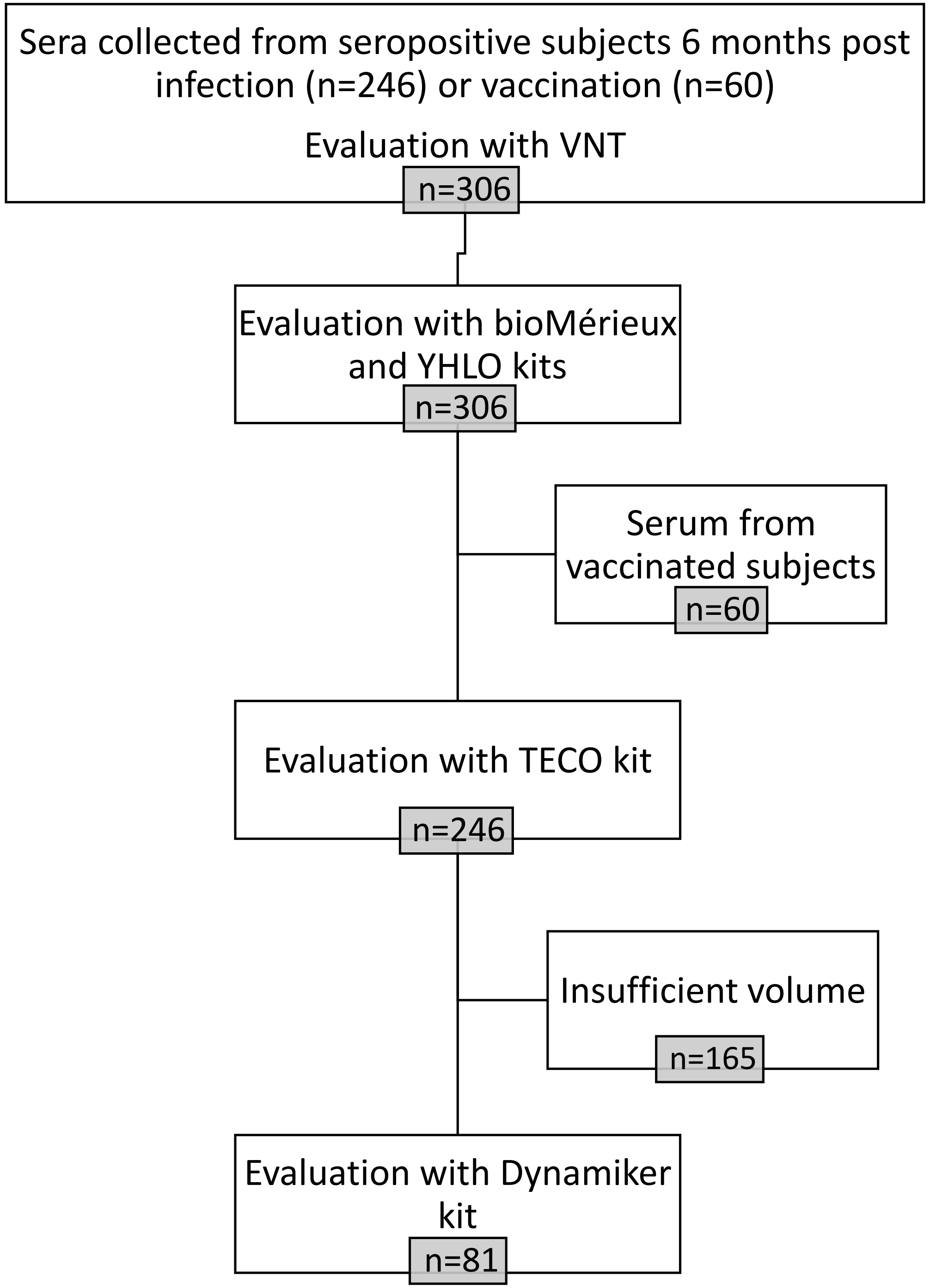
