## Supplementary materials and methods for "Evaluation of Commercial Anti-SARS-CoV-2 Neutralizing Antibody Assays in seropositive subjects"

**Samples treatment**

Serum samples were prepared from 5 mL of whole blood collected in BD Vacutainer^®^ Serum Separator Tubes II Advance (Becton Dickinson Diagnostics, le Pont-De-Claix, France). After collection, tubes were shaken gently and serum allowed to clot for a minimum 30 min at room temperature to obtain total coagulation, followed by centrifugation at 2,000 g for 10 min. Serum was stored at -80°C until serological assays were performed.

**Plaque of reduction neutralization test (PRNT_50_)**

All experiments were performed in a biosafety level 3 laboratory. Briefly, a 10-fold dilution of each serum specimen in culture medium (Dulbecco’s modified Eagle medium containing antibiotics and 2% fetal calf serum) was first heated for 30 min at 56°C to avoid complement-linked reduction of the viral activity. Serial 2-fold dilutions (tested in duplicate) of the serum specimens in culture medium were mixed at equal volume with the live SARS-CoV-2 virus. After gentle shaking and a contact of 30 min at room temperature in plastic microplates, 150 µL of the mix was transferred into 96-well microplates covered with Vero E6 cells. The plates were incubated at 37°C in a 5% CO2 atmosphere. The plates were read microscopically 5 days later when the cytopathic effect of the virus control reached 100 TCID50/150 µL (Tissue culture infective dose). Neutralization was recorded if more than 50% of the cells present in the well were preserved. The neutralizing titer was expressed as the inverse of the highest serum dilution that exhibited a 50% inhibition of infection;
